# Comparative effectiveness of sotrovimab and molnupiravir for preventing severe COVID-19 outcomes in non-hospitalised patients on kidney replacement therapy: observational cohort study using the OpenSAFELY-UKRR linked platform and SRR database

**DOI:** 10.1101/2022.12.02.22283049

**Authors:** The OpenSAFELY Collaborative, Bang Zheng, Jacqueline Campbell, Edward J Carr, John Tazare, Linda Nab, Viyaasan Mahalingasivam, Amir Mehrkar, Shalini Santhakumaran, Retha Steenkamp, Fiona Loud, Susan Lyon, Miranda Scanlon, William J Hulme, Amelia CA Green, Helen J Curtis, Louis Fisher, Edward Parker, Ben Goldacre, Ian Douglas, Stephen Evans, Brian MacKenna, Samira Bell, Laurie A Tomlinson, Dorothea Nitsch, The LH&W NCS (or CONVALESCENCE) Collaborative

## Abstract

**Background:** Patients on kidney replacement therapy (KRT; dialysis and kidney transplantation) are at the highest risk of severe outcomes from COVID-19. Due to limited inclusion of patients on KRT in clinical trials, information is limited on the effectiveness of sotrovimab (a neutralising monoclonal antibody). We sought to address this by comparing its effectiveness against molnupiravir (an antiviral) in preventing severe COVID-19 outcomes in non-hospitalised adults with symptomatic COVID-19.

**Methods:** With the approval of NHS England we used routine clinical data from 24 million patients in England linked to the UK Renal Registry (UKRR) to identify patients on KRT, and data on antiviral treatments, COVID-19 test results, hospitalisation events and death from the OpenSAFELY-TPP data resource. Cox proportional hazards models (stratified for region) were used to estimate hazard ratios of sotrovimab vs. molnupiravir with regards to COVID-19 related hospitalisation or deaths in the subsequent 28 days (as the primary outcome). Further analyses were conducted using propensity score weighting (adjusted for region) and to investigate robustness of results with regards to different time periods, missing data, and adjustment variables. We also conducted a complementary analysis using data from patients in the Scottish Renal Registry (SRR) treated with sotrovimab or molnupiravir, following similar analytical approaches.

**Results:** Among the 2367 renal patients treated with sotrovimab (n=1852) or molnupiravir (n=515) between December 16, 2021 and August 1, 2022 in England, 38 cases (1.6%) of COVID-19 related hospitalisations/deaths were observed during the 28 days of follow-up after treatment initiation, with 21 (1.1%) in the sotrovimab group and 17 (3.3%) in the molnupiravir group. In multiple-adjusted analysis sotrovimab was associated with substantially lower risk of 28-day COVID-19 related hospitalisation/death than treatment with molnupiravir (hazard ratio, HR=0.35, 95% CI: 0.17 to 0.71; P=0.004), with results remaining robust in sensitivity analyses. In the SRR cohort, there were 19 cases (1.9%) of COVID-19 related hospitalisations/deaths during the 28 days of follow-up after treatment initiation of sotrovimab (n=723) or molnupiravir (n=270). In multiple-adjusted analysis, sotrovimab showed a trend toward lower risk of 28-day COVID-19 related hospitalisation/death than treatment with molnupiravir (HR=0.39, 95% CI: 0.13 to 1.21; P=0.106). In both datasets, sotrovimab had no evidence of association with other hospitalisation/death compared with molnupiravir (HRs ranging from 0.73-1.29; P>0.05).

**Conclusions:** In routine care of non-hospitalised patients with COVID-19 on kidney replacement therapy, those who received sotrovimab had substantially lower risk of severe COVID-19 outcomes than those receiving molnupiravir.

## Background

People receiving kidney replacement therapy (KRT) are very vulnerable to severe outcomes from COVID-19.[1] This is due to effects from impaired kidney function, the underlying disease, and treatments affecting both underlying vulnerability to severe respiratory disease and vaccine response.[2] These biological factors intersect with reduced ability to shield due to needing to attend hospital for specialist care, particularly those people treated with in-centre haemodialysis (IC-HD).[3] Whilst vaccination has greatly improved the relative risk of severe outcomes for many of the originally identified vulnerable groups, such as older individuals,[4] it has offered modest gains for people receiving KRT.[5]

For both transplant and dialysis populations, there is substantial evidence of attenuated responses to vaccinations against pre-pandemic pathogens.[6] People receiving KRT were excluded from phase 3 SARS-CoV-2 vaccine trials [8-10] but in vitro studies of immunogenicity of AZD1222 and BNT162b2 showed reduced responses compared to people without kidney disease [11,12], particularly among people who were additionally immunosuppressed.[13,14] Hence, despite vaccination, many people receiving KRT remain at high risk of severe illness from SARS-CoV-2 infection, suggesting they may have substantial benefit from out-patient antiviral treatments. However, trials of these medications limited inclusion of immunosuppressed patients [15] or those on dialysis.[16] No current evidence from randomised trials exists among patients receiving KRT for sotrovimab, a neutralising monoclonal antibody (nMAb), although there is an ongoing platform trial for prophylaxis in vulnerable patient groups, including transplantation [NCT04870333]. Limited evidence is also available for molnupiravir, an oral antiviral, among people receiving KRT.[17] Nonetheless, in both England and Scotland antiviral medications were pragmatically recommended for people receiving KRT. These were deployed via dedicated treatment centres (COVID Medicine Delivery Units, CMDUs) in England and administered centrally via individual NHS health boards in Scotland, both established in December 2021 to provide timely antiviral treatment of vulnerable patients in the community.

We used two sources of high-quality routinely collected clinical data in England - the UK Renal Registry (UKRR) linked to the OpenSAFELY platform to enable comprehensive clinical data (including comorbidities, vaccination and infection history), and accurate identification of people receiving KRT, linked to treatment information about administered antiviral medication, to bridge this gap in knowledge for COVID-19 treatments. In addition, we conducted a complementary analysis following similar approaches using data from the Scottish Renal Registry (SRR). We sought to compare the effectiveness of sotrovimab vs. molnupiravir in preventing severe outcomes from COVID-19 in non-hospitalised symptomatic adult patients receiving KRT in England and Scotland.

## Methods

### Study population

#### OpenSAFELY-UKRR cohort

We included adults (≥18 years old) within the OpenSAFELY-TPP platform who were receiving KRT and had non-hospitalised treatment records for either sotrovimab or molnupiravir between December 16, 2021 and August 1, 2022. We focused on these two drugs because only a small number of infected patients on KRT were treated with Paxlovid (as severe kidney disease and immunosuppressive drugs used in kidney transplant patients are contraindications [18]), remdesivir, or casirivimab/imdevimab. During the early part of the study (from December 16, 2021 to February 10, 2022) there was relative clinical equipoise between sotrovimab and molnupiravir, with either agent recommended for treatment of symptomatic high risk patients in national guidance.[19]

We required patients to be registered at a GP surgery at the time of treatment initiation to allow for extraction of baseline and follow-up information. According to the eligibility criteria from NHS England for nMAb or antiviral treatment in the community, besides being in the renal disease cohort (one of the ten specified high-risk cohorts), the included patients were assumed to have SARS-CoV-2 infection confirmed by polymerase chain reaction (PCR) testing or lateral flow test, onset of COVID-19 symptoms within the last five days, have no signs of recovery and not require hospitalisation for COVID-19 or supplemental oxygen specifically for the management of COVID-19 symptoms before treatment initiation.

#### SRR cohort

All adults (≥18 years old) who were on KRT in Scotland who had a linked record for receiving either sotrovimab or molnupiravir between December 21, 2021 and August 31, 2022 were included. The eligibility criteria for nMAb or antiviral treatment in the community in Scotland was the same as for NHS England described above.

### Data sources

#### OpenSAFELY-UKRR cohort

All data were linked, stored and analysed securely within the OpenSAFELY platform: https://opensafely.org/. OpenSAFELY is a data analytics platform created by our team on behalf of NHS England to address urgent COVID-19 research questions. The dataset analysed within OpenSAFELY-TPP is based on 24 million people currently registered with GP surgeries using TPP SystmOne software. Data include pseudonymised data such as coded diagnoses, medications and physiological parameters. No free text data are included. All code is shared openly for review and re-use under MIT open license (https://github.com/opensafely/sotrovimab-and-molnupiravir). Detailed pseudonymised patient data is potentially re-identifiable and therefore not shared. Primary care records managed by the GP software provider TPP are securely linked to other similarly pseudonymised datasets, including the UKRR database, Office for National Statistics (ONS) mortality database, in-patient hospital spell records via Secondary Uses Service (SUS), national coronavirus testing records via the Second Generation Surveillance System (SGSS), and the COVID-19 therapeutics dataset, a patient-level dataset on nMAbs and antiviral treatments derived from Blueteq software that CMDUs use to notify NHS England of COVID-19 treatments. Patient-level vaccination status is available in the GP records directly via the National Immunisation Management System (NIMS).

The UKRR database contains data from patients under secondary renal care (advanced chronic kidney disease stages 4 and 5, dialysis, and kidney transplantation). In this study, we restricted our study population to those in the UKRR 2021 prevalence cohort (i.e., a prevalence cohort of patients alive and on KRT in December 2021).

#### SRR cohort

The Scottish Renal Registry is a national registry of all patients receiving KRT in Scotland (HD, peritoneal dialysis, and transplant). It collates data from all nine adult renal units in Scotland and 28 satellite HD units serving a population of 5.4 million with 100% unit and patient coverage. Data held by the registry include patient demographics, including historical postcodes (for calculating the Scottish Index of Multiple Deprivation [SIMD]),[20] full KRT history (for kidney failure), primary renal diagnosis (using ERA-EDTA codes). Data on SARS-CoV-2 testing were obtained from the Electronic Communication of Surveillance in Scotland, with date of test and result reported. Information on hospital admissions was obtained from the Scottish Morbidity Record and Rapid Preliminary Inpatient Data, and data on deaths were obtained from the National Records of Scotland. Details on vaccination type and date were obtained from the Turas Vaccination Management Tool, which holds all vaccination records in Scotland. Furthermore, data on treatment with sotrovimab or molnupiravir was obtained from information provided by the Health Boards to Public Health Scotland in addition to data obtained via the Hospital Electronic Prescribing and Medicines Administration (HEPMA) in the boards where this is available.

### Exposure

The exposure was treatment with sotrovimab or molnupiravir. In the OpenSAFELY-UKRR cohort, patients were excluded if they had treatment records of any other nMAbs or antivirals for COVID-19 before receiving sotrovimab or molnupiravir (n≤5). Patients with treatment records of both sotrovimab and molnupiravir were censored at the start date of the second treatment (n=8). In the SRR cohort, as the data linkage was only undertaken looking at sotrovimab or molnupiravir, we are unable to determine if any other antiviral treatments have been given prior to this.

### Outcomes

The primary outcome was COVID-19 related hospitalisation or COVID-19 related death within 28 days after treatment initiation. COVID-19 related hospitalisation was defined as hospital admission with COVID-19 as the primary diagnosis in the OpenSAFELY-UKRR cohort, and defined as emergency hospital admission with COVID-19 as the main condition in the SRR cohort. COVID-19 related death was defined as COVID-19 being the underlying/contributing cause of death in death certificates in both cohorts (based on ICD-10 codes U07.1 and U07.2).

Secondary outcomes were 28-day all-cause hospital admission or death, and 60-day COVID-19 related hospitalisation/death. In the OpenSAFELY-UKRR cohort, to exclude events where patients were admitted in order to receive sotrovimab or other planned/regular treatment (e.g., dialysis), we did not count admissions coded as “elective day case admission” or “regular admission” in SUS or day cases detected by the same admission and discharge dates as hospitalisation events (**Supplementary Table 1**). Similarly, in the SRR cohort, only emergency hospital admissions with the length of hospital stay greater than 0 were counted as outcome events.

### Covariates

#### OpenSAFELY-UKRR cohort

The following covariates were extracted at baseline: KRT modality (dialysis or kidney transplantation), years since KRT start date, age, sex, NHS region, ethnicity (White or non-White), Index of Multiple Deprivation (IMD, as quintiles derived from the patient’s postcode at lower super output area level to reflect socio-economic status), rural-urban classification (derived from patient’s postcode), calendar date (to account for secular trends of prescription and incidence rate of COVID-19 outcomes), COVID-19 vaccination status (unvaccinated/one/two vaccinations, or three or more vaccinations), positive test date for SARS-CoV-2 infection (PCR or lateral flow test, as a proxy for symptom onset date), body mass index (BMI, the most recent record within 10 years; <25 kg/m^2^, 25 - <30 kg/m^2^, ≥30 kg/m^2^), high-risk cohort categories other than renal disease or kidney transplantation (Down syndrome, a solid cancer, a haematological disease or stem cell transplant, liver disease, immune-mediated inflammatory disorders, immunosuppression, HIV/AIDS, or rare neurological conditions; allowing multiple categories per patient), other comorbidities (diabetes, hypertension, chronic cardiac disease, chronic respiratory disease, learning disabilities, severe mental illness), and care home residency and housebound status.

#### SRR cohort

The following covariates were extracted at baseline: KRT modality (dialysis or kidney transplantation), years since KRT start date, age, sex, Scottish IMD (based on patient postcode, as quintiles with one corresponding to most deprived and five corresponding to least deprived),[20] calendar date of treatment initiation, COVID-19 vaccination status (unvaccinated/one/two vaccinations, or three or more vaccinations), positive test date for SARS-CoV-2 infection (PCR or lateral flow test) and primary renal diagnosis (PRD).

### Statistical analyses

#### OpenSAFELY-UKRR cohort

Distributions of baseline characteristics were compared between the two treatment groups. Follow-up time of individual patients was calculated from the recorded treatment initiation date, until the outcome event date, 28 days after treatment initiation, initiation of a second nMAb/antiviral treatment, death, patient deregistration date, or the study end date (September 30, 2022), whichever occurred first.

Risks of 28-day COVID-19 related hospitalisation/death were compared between the two groups using Cox proportional hazards models, with time since treatment as the time scale. The Cox models were stratified by NHS region to account for geographic heterogeneity in baseline hazards, with sequential adjustment for other baseline covariates. Model 1 was adjusted for age and sex; Model 2 additionally adjusted for high-risk cohort categories (except Down syndrome, liver disease, HIV/AIDS and rare neurological conditions due to low counts), KRT modality and years since KRT start date; Model 3 further adjusted for ethnicity, IMD quintiles, vaccination status, calendar date (with restricted cubic splines to account for non-linear effect); and Model 4 additionally adjusted for BMI category, diabetes, hypertension, chronic cardiac and respiratory diseases. Missing values of covariates were treated as separate categories to retain sample size. The proportional hazards assumption was tested based on the scaled Schoenfeld residuals.

As an alternative approach, we adopted the propensity score weighting (PSW) method to account for confounding bias. The covariates were balanced between the two drug groups through the average treatment effect (ATE) weighting scheme based on the estimated propensity scores. Balance check of baseline covariates after weighting was conducted using standardised mean differences between groups (with threshold of <0.10 as the indicator of well-balanced). Robust variance estimators were used in the weighted Cox models.

Similar analytical procedures were used for secondary outcomes. In addition, we explored whether the following factors could modify the observed comparative effectiveness: time period with different dominant variants (December 16, 2021 to February 15, 2022 for BA.1, February 16-May 31 for BA.2, June 1-August 1 for BA.4/BA.5),[21] KRT modality (dialysis or kidney transplantation), BMI categories (≥30 vs. <30 kg/m^2^), presence of diabetes, hypertension, chronic cardiac diseases or chronic respiratory diseases, days between test positive and treatment initiation (<3 vs. 3-5 days), age group (<60 vs. ≥60 years), sex and ethnicity (White vs. non-White). Effect modification by each covariate was tested by adding the corresponding interaction term in the stratified Cox model.

Additional sensitivity analyses based on the stratified Cox models were conducted, including (1) using complete case analysis or Multiple Imputation by Chained Equations to deal with missing values in covariates; (2) using Cox models with calendar date as the underlying time scale to further account for temporal trends (and circulating variants); (3) additionally adjusting for time between test positive and treatment initiation, and time between last vaccination date and treatment initiation; (4) additionally adjusting for rural-urban classification, and other comorbidities and factors that might have influenced clinician’s choice of therapy through the patient’s ability to travel to hospital for an infusion (learning disabilities, severe mental illness, care home residency or housebound status); (5) using restricted cubic splines for age to further control for potential non-linear age effect; (6) excluding patients with treatment records of both sotrovimab and molnupiravir, or with treatment records of casirivimab/imdevimab, Paxlovid, or remdesivir; (7) excluding patients who did not have a positive SARS-CoV-2 test record before treatment or initiated treatment after 5 days since positive SARS-CoV-2 test; (8) creating a 1-day or 2-day lag in the follow-up start date to account for potential delays in drug administration; (9) conducting a cause-specific analysis for the 28-day COVID-19 related hospitalisation/death vs. other hospitalisation/death.

#### SRR cohort

Similar statistical analyses were conducted in the SRR cohort, except where there was no relevant covariate information.

### Software and reproducibility

In the UKRR analysis, data management was performed using Python, with analysis carried out using Stata 16.1. Code for data management and analysis, as well as codelists, are archived online (https://github.com/opensafely/sotrovimab-and-molnupiravir). For the SRR analysis, data management and analyses were performed by a Public Health Scotland analyst within Public Health Scotland using R studio v3.6.1.

### Patient and public involvement

We have developed a publicly available website https://opensafely.org/ through which we invite any patient or member of the public to make contact regarding this study or the broader OpenSAFELY project. Patient representatives including those representing the UKRR patient council and KidneyCare UK have actively contributed to the presentation of these results and are co-authors on this paper.

## Results

### OpenSAFELY-UKRR cohort

#### Patient characteristics

Between December 16, 2021 and August 1, 2022, a total of 2367 non-hospitalised COVID-19 patients on KRT were treated with sotrovimab (n=1852) or molnupiravir (n=515). The mean age of these patients was 55.9 (SD=14.6) years; 43.5% were female, 85.4% were White and 92.6% had three or more COVID-19 vaccinations. In the whole treated population, 69.6% were kidney transplant recipients and 30.4% were on dialysis. Among these 81.8% of dialysis patients and 76.7% of transplant patients were treated with sotrovimab. Baseline characteristics were similar between the groups receiving different treatments (**Table 1**) but the sotrovimab group had a lower proportion of kidney transplant recipients (68.3% vs. 74.6%) and a higher proportion of patients with chronic cardiac disease (27.3% vs. 21.6%). There were also some geographic variations in the prescription of these two drugs and greater use of molnupiravir earlier during the study period.

**Table 1.**
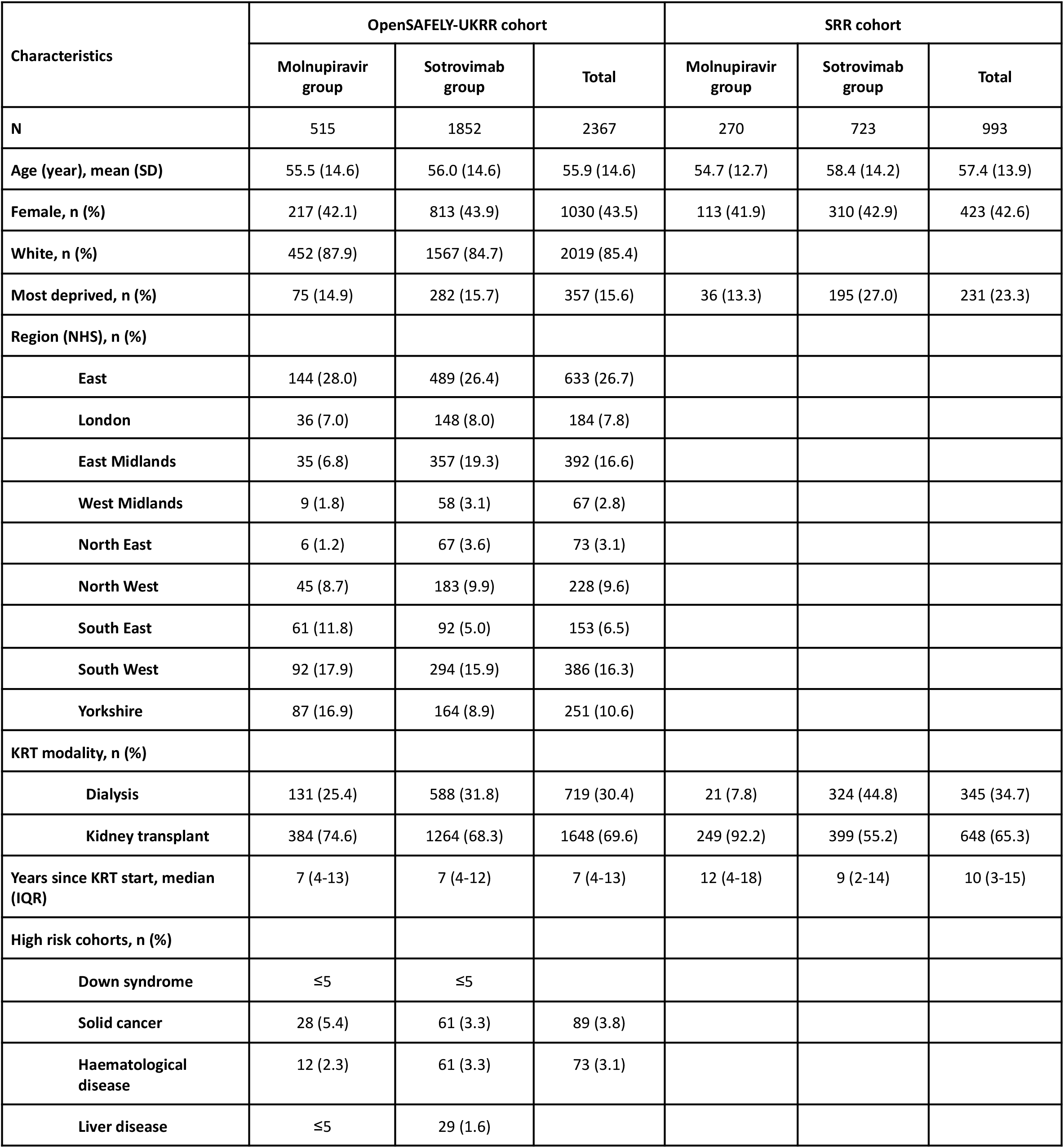
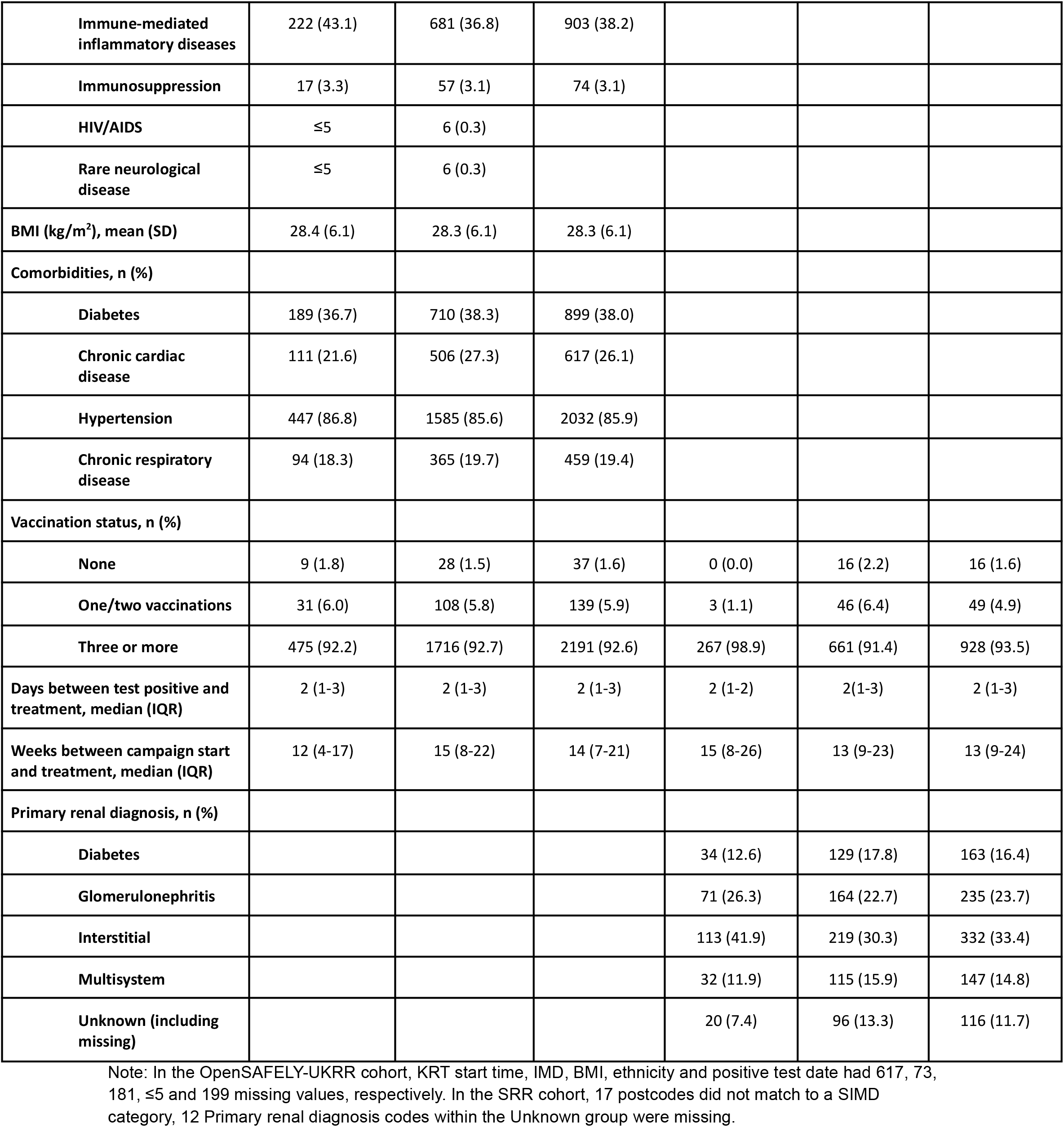
Baseline characteristics of patients on KRT receiving molnupiravir or sotrovimab.

| Characteristics | OpenSAFELY-UKRR cohort |  |  | SRR cohort |  |  |
| --- | --- | --- | --- | --- | --- | --- |
|  | Molnupiravir group | Sotrovimab group | Total | Molnupiravir group | Sotrovimab group | Total |
| **N** | 515 | 1852 | 2367 | 270 | 723 | 993 |
| **Age (year), mean (SD)** | 55.5 (14.6) | 56.0 (14.6) | 55.9 (14.6) | 54.7 (12.7) | 58.4 (14.2) | 57.4 (13.9) |
| **Female, n (%)** | 217 (42.1) | 813 (43.9) | 1030 (43.5) | 113 (41.9) | 310 (42.9) | 423 (42.6) |
| **White, n (%)** | 452 (87.9) | 1567 (84.7) | 2019 (85.4) |  |  |  |
| **Most deprived, n (%)** | 75 (14.9) | 282 (15.7) | 357 (15.6) | 36 (13.3) | 195 (27.0) | 231 (23.3) |
| **Region (NHS), n (%)** |  |  |  |  |  |  |
| **East** | 144 (28.0) | 489 (26.4) | 633 (26.7) |  |  |  |
| **London** | 36 (7.0) | 148 (8.0) | 184 (7.8) |  |  |  |
| **East Midlands** | 35 (6.8) | 357 (19.3) | 392 (16.6) |  |  |  |
| **West Midlands** | 9 (1.8) | 58 (3.1) | 67 (2.8) |  |  |  |
| **North East** | 6 (1.2) | 67 (3.6) | 73 (3.1) |  |  |  |
| **North West** | 45 (8.7) | 183 (9.9) | 228 (9.6) |  |  |  |
| **South East** | 61 (11.8) | 92 (5.0) | 153 (6.5) |  |  |  |
| **South West** | 92 (17.9) | 294 (15.9) | 386 (16.3) |  |  |  |
| **Yorkshire** | 87 (16.9) | 164 (8.9) | 251 (10.6) |  |  |  |
| **KRT modality, n (%)** |  |  |  |  |  |  |
| **Dialysis** | 131 (25.4) | 588 (31.8) | 719 (30.4) | 21 (7.8) | 324 (44.8) | 345 (34.7) |
| **Kidney transplant** | 384 (74.6) | 1264 (68.3) | 1648 (69.6) | 249 (92.2) | 399 (55.2) | 648 (65.3) |
| **Years since KRT start, median (IQR)** | 7 (4-13) | 7 (4-12) | 7 (4-13) | 12 (4-18) | 9 (2-14) | 10 (3-15) |
| **High risk cohorts, n (%)** |  |  |  |  |  |  |
| **Down syndrome** | ≤5 | ≤5 |  |  |  |  |
| **Solid cancer** | 28 (5.4) | 61 (3.3) | 89 (3.8) |  |  |  |
| **Haematological disease** | 12 (2.3) | 61 (3.3) | 73 (3.1) |  |  |  |
| **Liver disease** | ≤5 | 29 (1.6) |  |  |  |  |
| **Immune-mediated inflammatory diseases** | 222 (43.1) | 681 (36.8) | 903 (38.2) |  |  |  |
| **Immunosuppression** | 17 (3.3) | 57 (3.1) | 74 (3.1) |  |  |  |
| **HIV/AIDS** | ≤5 | 6 (0.3) |  |  |  |  |
| **Rare neurological disease** | ≤5 | 6 (0.3) |  |  |  |  |
| **BMI (kg/m<sup>2</sup>), mean (SD)** | 28.4 (6.1) | 28.3 (6.1) | 28.3 (6.1) |  |  |  |
| **Comorbidities, n (%)** |  |  |  |  |  |  |
| **Diabetes** | 189 (36.7) | 710 (38.3) | 899 (38.0) |  |  |  |
| **Chronic cardiac disease** | 111 (21.6) | 506 (27.3) | 617 (26.1) |  |  |  |
| **Hypertension** | 447 (86.8) | 1585 (85.6) | 2032 (85.9) |  |  |  |
| **Chronic respiratory disease** | 94 (18.3) | 365 (19.7) | 459 (19.4) |  |  |  |
| **Vaccination status, n (%)** |  |  |  |  |  |  |
| **None** | 9 (1.8) | 28 (1.5) | 37 (1.6) | 0 (0.0) | 16 (2.2) | 16 (1.6) |
| **One/two vaccinations** | 31 (6.0) | 108 (5.8) | 139 (5.9) | 3 (1.1) | 46 (6.4) | 49 (4.9) |
| **Three or more** | 475 (92.2) | 1716 (92.7) | 2191 (92.6) | 267 (98.9) | 661 (91.4) | 928 (93.5) |
| **Days between test positive and treatment, median (IQR)** | 2 (1-3) | 2 (1-3) | 2 (1-3) | 2 (1-2) | 2(1-3) | 2 (1-3) |
| **Weeks between campaign start and treatment, median (IQR)** | 12 (4-17) | 15 (8-22) | 14 (7-21) | 15 (8-26) | 13 (9-23) | 13 (9-24) |
| **Primary renal diagnosis, n (%)** |  |  |  |  |  |  |
| **Diabetes** |  |  |  | 34 (12.6) | 129 (17.8) | 163 (16.4) |
| **Glomerulonephritis** |  |  |  | 71 (26.3) | 164 (22.7) | 235 (23.7) |
| **Interstitial** |  |  |  | 113 (41.9) | 219 (30.3) | 332 (33.4) |
| **Multisystem** |  |  |  | 32 (11.9) | 115 (15.9) | 147 (14.8) |
| **Unknown (including missing)** |  |  |  | 20 (7.4) | 96 (13.3) | 116 (11.7) |

Note: In the OpenSAFELY-UKRR cohort, KRT start time, IMD, BMI, ethnicity and positive test date had 617, 73, 181, ≤5 and 199 missing values, respectively. In the SRR cohort, 17 postcodes did not match to a SIMD category, 12 Primary renal diagnosis codes within the Unknown group were missing.

#### Comparative effectiveness for the outcome events

Among the 2367 renal patients treated with sotrovimab or molnupiravir, 38 cases (1.6%) of COVID-19 related hospitalisations/deaths were observed during the 28 days of follow-up after treatment initiation, with 21 (1.1%) in the sotrovimab group and 17 (3.3%) in the molnupiravir group; the number of COVID-19 related deaths were ≤5 in both groups.

Results of stratified Cox regression showed that, after adjusting for demographic variables, KRT modality and duration, high-risk cohort categories, vaccination status, calendar date, BMI category and other comorbidities, treatment with sotrovimab was associated with substantially lower risk of 28-day COVID-19 related hospitalisation/death than treatment with molnupiravir (hazard ratio, HR=0.35, 95% CI: 0.17 to 0.71; P=0.004). Consistent results favouring sotrovimab over molnupiravir were obtained from propensity score weighted Cox models (Model 4: HR=0.39, 95% CI: 0.19 to 0.80; P=0.010), following confirmation of successful balance of baseline covariates between groups in the weighted sample (**Supplementary Table 2**). The magnitude of HRs was stable during the sequential covariate adjustment process (ranging from 0.32-0.35 across different models; **Figure 1**). No violation of the proportional hazards assumption was detected in any model (P>0.10).

**Figure 1.**
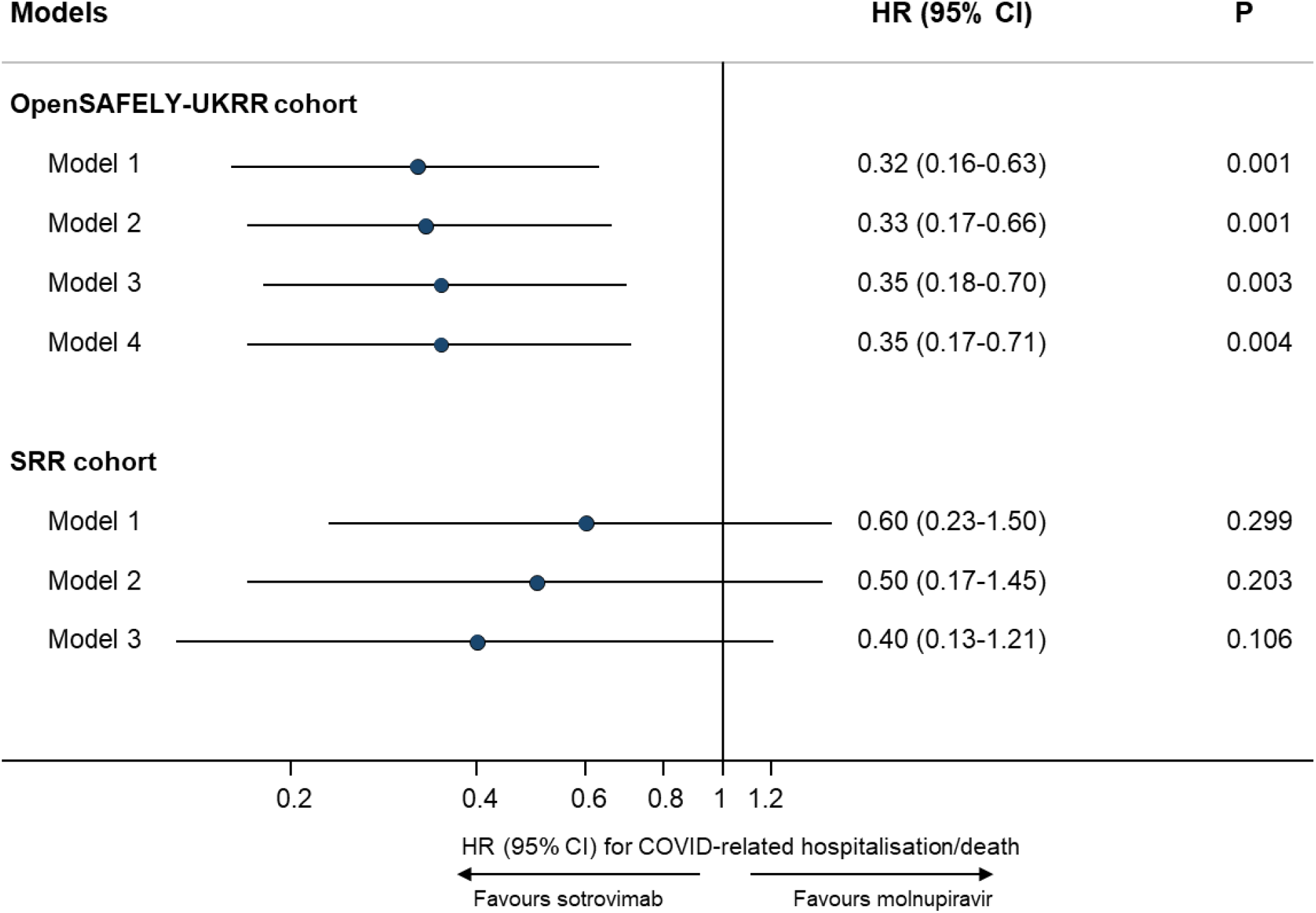
Comparing risk of 28-day COVID-19 related hospitalisation/death between sotrovimab vs. molnupiravir in two cohorts. Note: HR=hazard ratio; CI=confidence interval. In the OpenSAFELY-UKRR cohort, Model 1 adjusted for age and sex; Model 2 additional adjusted for high risk cohort categories, KRT modality and duration; Model 3 further adjusted for ethnicity, IMD quintiles, vaccination status, calendar date; and Model 4 additionally adjusted for BMI category, diabetes, hypertension, chronic cardiac and respiratory diseases. In the SRR cohort, Model 1 adjusted for age and sex; Model 2 additionally adjusted for modality, PRD Group and KRT duration; Model 3 additionally adjusted for SIMD, vaccination status and calendar time.

For the secondary outcomes, the analysis of 60-day COVID-19 related events revealed similar results in favour of sotrovimab (HRs ranging from 0.33-0.36; P<0.05). For all-cause hospitalisations/deaths, 163 cases (6.9%) were observed during the 28 days of follow-up after treatment initiation (117 [6.4%] in the sotrovimab group and 46 [9.0%] in the molnupiravir group). Results of stratified Cox regression showed a lower risk in the sotrovimab group than in the molnupiravir group (HRs ranging from 0.60-0.65 in Models 1-4; P<0.05; **Table 2**).

**Table 2.** Comparing risks of non-COVID-specific outcomes between sotrovimab vs. molnupiravir in two cohorts.

| Outcomes | OpenSAFELY-UKRR cohort |  |  | SRR cohort |  |  |
| --- | --- | --- | --- | --- | --- | --- |
|  | N/Events | HR (95% CI) for sotrovimab (ref=molnupiravir) | P | N/Events | HR (95% CI) for sotrovimab (ref=molnupiravir) | P |
| **28-day All-cause hospitalisation/death** | 2350/163 |  |  | 993/75 |  |  |
| **Model 1** |  | 0.65 (0.46-0.92) | 0.016 |  | 1.04 (0.61-1.76) | 0.879 |
| **Model 2** |  | 0.63 (0.44-0.89) | 0.010 |  | 0.80 (0.45-1.43) | 0.455 |
| **Model 3** |  | 0.62 (0.43-0.89) | 0.009 |  | 0.71 (0.39-1.29) | 0.273 |
| **Model 4** |  | 0.60 (0.41-0.85) | 0.004 |  |  |  |
| **28-day Other-cause hospitalisation/death** | 2350/130 |  |  | 993/56 |  |  |
| **Model 1** |  | 0.85 (0.56-1.29) | 0.441 |  | 1.29 (0.68-2.40) | 0.426 |
| **Model 2** |  | 0.79 (0.52-1.21) | 0.276 |  | 0.97 (0.47-1.95) | 0.934 |
| **Model 3** |  | 0.76 (0.49-1.16) | 0.205 |  | 0.90 (0.44-1.84) | 0.776 |
| **Model 4** |  | 0.73 (0.48-1.12) | 0.151 |  |  |  |

Note: HR=hazard ratio; CI=confidence interval. In the OpenSAFELY-UKRR cohort, Model 1 adjusted for age and sex; Model 2 additional adjusted for high risk cohort categories, KRT modality and duration; Model 3 further adjusted for ethnicity, IMD quintiles, vaccination status, calendar date; and Model 4 additionally adjusted for BMI category, diabetes, hypertension, chronic cardiac and respiratory diseases. In the SRR cohort, Model 1 adjusted for age and sex; Model 2 additionally adjusted for modality, PRD Group and KRT duration; Model 3 additionally adjusted for SIMD, vaccination status and calendar time.

#### Sensitivity analyses and tests for effect modification

Results of sensitivity analyses were consistent with the main findings (**Supplementary Table 3**). Among patients included in the cause-specific analysis (n=2350), 33 had COVID-19 related hospitalisation/death and 130 had other hospitalisation/death events within 28 days after treatment initiation. The cause-specific Cox model showed that, unlike COVID-related outcomes, there was no evidence of an association of sotrovimab with other hospitalisation/death compared with molnupiravir (HRs ranging from 0.73-0.85 in Models 1-4; P>0.05; **Table 2**) despite a greater number of events compared to the primary outcome.

No substantial effect modification was observed for time period of treatment, KRT modality, presence of obesity, diabetes, hypertension, chronic cardiac diseases or chronic respiratory diseases, time since test positive, age group, sex or ethnicity (P for interaction>0.10; **Figure 2**).

**Figure 2.**
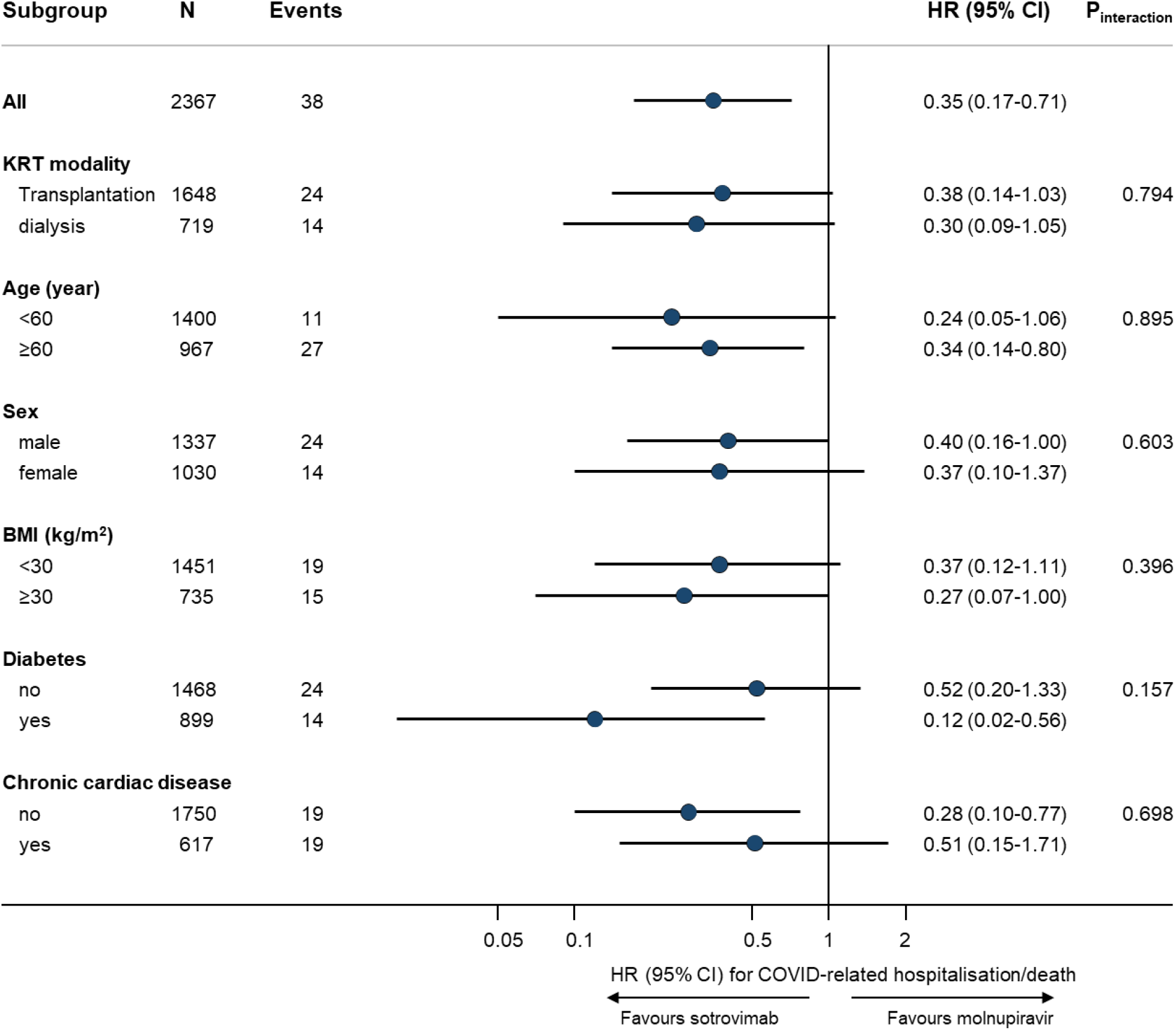
Subgroup analysis of sotrovimab vs. molnupiravir in association with risk of 28-day COVID-19 related hospitalisation/death (OpenSAFELY-UKRR cohort). Note: HR=hazard ratio; CI=confidence interval; BMI=body mass index. Subgroup analyses were based on the fully-adjusted stratified Cox model (Model 4). P for interaction between drug group and each of the following variables was: time period 16/2/2022-31/5/2022 (0.577), time period 1/6/2022-1/8/2022 (0.640), hypertension (0.286), chronic respiratory diseases (0.449), days between test positive and treatment initiation (0.377), and White ethnicity (0.379), respectively; no analyses within each level of these variables were done because of lack of sample size or outcome events within the subset of population.

### SRR cohort

Between December 21, 2021 and August 31, 2022, a total of 993 non-hospitalised COVID-19 patients on KRT were treated with sotrovimab (n=723) or molnupiravir (n=270). The mean age of these patients was 57.4 (SD=13.9) years; 42.6% were female and 93.5% had three or more COVID-19 vaccinations; 65.3% were kidney transplant recipients and 34.7% were on dialysis. Compared to the molnupiravir group, the sotrovimab group had a lower proportion of kidney transplant recipients (55.2% vs. 92.2%; **Table 1**).

During the 28 days of follow-up after treatment initiation, 19 cases (1.9%) of COVID-19 related hospitalisations/deaths were observed, with 12 (1.7%) in the sotrovimab group and 7 (2.6%) in the molnupiravir group. There were 6 COVID-19 related deaths in the sotrovimab group and 5 in the molnupiravir group.

Results of Cox regression showed that, after adjusting for age, sex, modality, primary renal diagnosis, SIMD, vaccination status, KRT duration and calendar date, treatment with sotrovimab was consistent with lower risk of 28-day COVID-19 related hospitalisation/death than treatment with molnupiravir although confidence intervals were broad and crossed the null (HR=0.39, 95% CI: 0.13 to 1.21; P=0.106; **Figure 1**). There was no substantial difference between sotrovimab and molnupiravir in the risk of all-cause hospitalisation/death (HRs ranging from 0.71-1.04 in Models 1-3; P>0.05) or other hospitalisation/death (HRs ranging from 0.90-1.29 in Models 1-3; P>0.05; **Table 2**).

## Discussion

### Summary

Our analysis shows that among people receiving KRT, treatment with sotrovimab is associated with a lower risk of severe outcomes from COVID-19 infection compared with molnupiravir during the Omicron wave in England in 2021-2022. We used a range of analytic methods to examine robustness of results, and were able to carry out extensive adjustments for confounding given the availability of granular multisource real-world data. Analyses in an independent dataset from the Scottish Renal Registry showed consistent effect estimates.

### Strengths and weaknesses

This study used two validated KRT populations from 2021 reported by all kidney care centres in England and Scotland at the start of the Omicron outbreak in two independent analyses which gave broadly similar results. This is the first time analyses from the two independent Renal Registries, both recognised as high-quality and complete data sources, have been combined. The English data were combined with real-time multisource data from the OpenSAFELY resource which allowed extensive adjustment for confounding. The Scottish data had less statistical power and less granular variables for confounding adjustment, and yielded more unstable point estimates across different statistical approaches (e.g., HR for 28-day COVID-19 related outcomes being 0.39 in Cox regression and 0.78 in propensity score analysis). Of note, in the English data detailed adjustment for confounding did not materially change the results.

While all patients in this analysis were eligible for antiviral treatment, it is possible that people with high health literacy or better baseline health status were more able to navigate timely access to antiviral therapy within their local health systems. While the study results could be due to such people being more likely to be treated with sotrovimab, baseline characteristics are very similar between people receiving the two treatments. If people perceived to be at highest risk of severe outcomes such as those receiving intensive immunosuppression or with underlying haematological disease were more likely to be treated with sotrovimab, this would be anticipated to bias the results in favour of molnupiravir, the converse of what is seen here. Similarly, while there will have been variation between dialysis units to the extent to which people with mild/moderate symptoms of COVID-19 will have been referred for antiviral therapy, this is unlikely to have affected the type of chosen antiviral as treatment decisions were made by dedicated antiviral treatment units.

There are regional variations in terms of immune priming and survivorship bias in the KRT population because the pandemic has affected different parts of the country in different ways.[22] Similarly, there may be regional variations in how referral pathways operated for patients to receive antiviral treatment during the Omicron pandemic, which could underlie the marked regional variation of antiviral use in our data. To account for these differences we stratified UKRR-OpenSAFELY data analyses by English region and adjusted for region in propensity score analyses.

Despite the granular data on underlying health status, the possibility of residual confounding cannot be ruled out in this real-world observational study. In February 2022 prescribing guidelines changed and molnupiravir was de-prioritised as a third-line treatment option,[18] which reduces therapeutic equipoise and may make these two treatment groups less comparable. A pointer towards potential residual confounding may be the association between treatment and all-cause hospitalisation and death, which was not observed in the general population.[23] However, in this KRT population with high levels of comorbidity and frailty, it is possible that more effective treatment of COVID-19 reduced incidence of other outcomes to an observable extent. Overall, given the size of the observed protective effect of sotrovimab and its robustness across multiple sensitivity analyses, residual confounding would have to be substantial to fully explain the findings. In addition, consistent findings in independent validation in the SRR where sources of bias and treatment pathways differed adds further robustness to the analysis.

### Findings in Context

Whilst the findings are in line with the COMET-ICE trial [15,24] in terms of showing a benefit for sotrovimab when compared to placebo (adjusted relative risk=0.21; absolute risk difference=-4.53%, 95% CI: −6.70% to −2.37%), there are important differences. The COMET-ICE trial included unvaccinated patients infected before the Omicron wave and excluded patients with immunosuppression; there were only 13 CKD patients amongst the 1057 patients who were randomised to either sotrovimab or placebo.

The phase 3 component of MOVe-OUT trial [16] was a double-blind RCT for molnupiravir in unvaccinated adults with mild-to-moderate COVID-19 and at least one risk factor for severe illness which showed a weaker effect (relative risk=0.70; absolute risk difference=-3.0%, 95% CI: −5.9% to −0.1%). There were more patients with CKD included (84 out of 1433 patients randomised to either molnupiravir or placebo), but the protocol excluded patients with an eGFR<30ml/min/1.73m^2^ or on dialysis. Another large-scale pragmatic trial for molnupiravir, the UK PANORAMIC trial (including only 480 kidney patients), showed that molnupiravir did not reduce risk of hospitalisations/deaths among high-risk vaccinated adults with COVID-19 in the community (25,000 participants, adjusted odds ratio=1.06, 95% Bayesian credible interval: 0.80 to 1.40).[17]

### Policy Implications and Interpretation

Currently, the living WHO guideline [25] makes weak recommendation in favour of molnupiravir and strong recommendation against sotrovimab for the treatment of non-severe COVID-19 patients. In addition, a draft technology appraisal by the National Institute for Health and Care Excellence does not recommend use of either agent.[26] In the latest version of NHS guideline on COVID-19 therapies for non-hospitalised patients, molnupiravir remains as the third-line option, but sotrovimab is only to be considered by exception where the available antivirals are contraindicated or determined to be unsuitable following multi-disciplinary team assessment.[27] However, there remains significant debate over the validity of the in vitro data that led to those decisions on sotrovimab.[28] Other recommended treatments such as nirmatrelvir/ritonavir are contraindicated in the KRT population. In this context, our analysis shows that for the KRT population treatment of COVID-19 with sotrovimab appears to be substantially more beneficial than molnupiravir.

### Future Research

Despite using English and Scottish data on patients on KRT, the sample size was still limited in terms of being able to detect rare side effects. The relative efficacy of sotrovimab vs molnupiravir may change when patients on KRT are exposed to other COVID-19 variants.

### Summary

People receiving KRT were largely under-represented or excluded from randomised trials of COVID-19 treatments. In routine care of non-hospitalised adults receiving KRT, sotrovimab was associated with a substantially lower risk of severe COVID-19 outcomes compared to molnupiravir.

## Data Availability

All data were linked, stored and analysed securely within the OpenSAFELY platform: https://opensafely.org/. All code is shared openly for review and re-use under MIT open license (https://github.com/opensafely/sotrovimab-and-molnupiravir). Detailed pseudonymised patient data is potentially re-identifiable and therefore not shared.

## Tables and Figures

**Supplementary Table 1.** Detected day case admissions that were not counted as hospitalisation events (OpenSAFELY-UKRR cohort).

| Admissions of which the admission and discharge dates were the same | Number of events |  |
| --- | --- | --- |
|  | Molnupiravir | Sotrovimab |
| Detected day cases on Day 0 (i.e., the treatment start date) | 23 | 105 |
| Those with MAb procedure on Day 0 | 6 | 94 |
| Detected day cases on Day 1 | ≤5 | 11 |
| Those with MAb procedure on Day 1 | ≤5 | 6 |
| Detected day cases on or after Day 2 | ≤5 | ≤5 |
| Those with MAb procedure on or after Day 2 | ≤5 | ≤5 |

Note: The following admissions were not counted as hospitalisation events: (1) those recorded as “elective day case admission” or “regular admission” in the patient classification field of SUS dataset, because such coding clearly indicates that the patient was admitted for a planned procedure or regular treatment (thus cannot represent severe COVID-19 outcomes); or (2) day cases that we additionally detected with the admission and discharge dates being the same. As shown in this table, most of those additionally detected day case admissions were for monoclonal antibody infusion procedure (i.e., admitted for receiving sotrovimab). Monoclonal antibody procedures were identified based on OPCS codes (X891, X892).

**Supplementary Table 2.** Results of propensity score weighting analyses in both cohorts.

| Outcome | OpenSAFELY-UKRR cohort |  |  | SRR cohort |  |  |
| --- | --- | --- | --- | --- | --- | --- |
|  | N/Events | HR (95% CI) for sotrovimab (ref=molnupiravir) | p | N/Events | HR (95% CI) for sotrovimab (ref=molnupiravir) | p |
| 28-day COVID-19 related hospitalisation/death | 2367/38 |  |  | 993/19 |  |  |
| Model 1 |  | 0.33 (0.16-0.64) | 0.001 |  | 0.61 (0.24-1.55) | 0.296 |
| Model 2 |  | 0.35 (0.18-0.70) | 0.003 |  | 0.87 (0.33-2.20) | 0.784 |
| Model 3 |  | 0.41 (0.20-0.83) | 0.013 |  | 0.78 (0.29-2.04) | 0.611 |
| Model 4 |  | 0.39 (0.19-0.80) | 0.010 |  |  |  |

Note: In the balance check of both cohorts, standardised mean differences of all covariates between groups after weighting were <0.1.

**Supplementary Table 3.** Sensitivity analysis of sotrovimab vs. molnupiravir in association with risk of 28-day COVID-19 related hospitalisation/death (OpenSAFELY-UKRR cohort).

| Sensitivity analyses | N | Events | HR (95% CI) for sotrovimab (ref=molnupiravir) | P |
| --- | --- | --- | --- | --- |
| Main analysis (for comparison purpose) | 2367 | 38 | 0.35 (0.17-0.71) | 0.004 |
| Additionally adjusting for days between test positive and treatment initiation, and months between last vaccination date and treatment initiation | 2367 | 38 | 0.35 (0.17-0.72) | 0.004 |
| Using Cox models with calendar date as the underlying time scale | 2367 | 38 | 0.38 (0.17-0.82) | 0.015 |
| Using restricted cubic splines for age | 2367 | 38 | 0.35 (0.17-0.71) | 0.004 |
| Additionally adjusting for rural-urban classification, comorbidities, care home and housebound status | 2367 | 38 | 0.27 (0.13-0.56) | <0.001 |
| Excluding patients with treatment records of both sotrovimab and molnupiravir, or any other therapies | 2359 | redacted | 0.35 (0.17-0.71) | 0.004 |
| Excluding patients without positive test record before treatment or initiated treatment after 5 days since positive test | 2032 | 30 | 0.43 (0.19-0.96) | 0.040 |
| Creating a 1-day lag in the follow-up start date | 2356 | redacted | 0.29 (0.14-0.62) | 0.001 |
| Creating a 2-day lag in the follow-up start date | 2351 | 31 | 0.33 (0.15-0.73) | 0.006 |
| Multiple imputation for covariates | 2367 | 38 | 0.39 (0.19-0.79) | 0.009 |
| Complete-case analysis | 1580 | 24 | 0.38 (0.15-0.96) | 0.040 |

Note: HR=hazard ratio; CI=confidence interval. Sensitivity analyses were based on the fully-adjusted stratified Cox model (Model 4).

## Abbreviations

COVID-19: Coronavirus Disease 2019
HR: hazard ratio
KRT: kidney replacement therapy
UKRR: UK Renal Registry
SRR: Scottish Renal Registry
95%: CI 95% confidence interval
PCR: polymerase chain reaction
IMD: Index of Multiple Deprivation
PSW: propensity score weighting

## Administrative

## Acknowledgements

We are very grateful for all the support received from the TPP Technical Operations team throughout this work, and for generous assistance from the information governance and database teams at NHS England / NHSX. We are also grateful to the UKRR Patient Council and KidneyCare UK who supported the data linkage to OpenSAFELY, and to colleagues at Public Health Scotland (Chrissie Watters, Euan Proud and Marion Bennie) and the SRR Steering group (Alison Almond, Katharine Buck, Wan Wong, Nicola Joss, Michaela Petrie, Shona Methven, Elaine Spalding and Peter Thomson).

## Information governance and ethical approval

OpenSAFELY: NHS England is the data controller; TPP is the data processor; and the researchers on OpenSAFELY are acting with the approval of NHS England. This implementation of OpenSAFELY is hosted within the TPP environment which is accredited to the ISO 27001 information security standard and is NHS IG Toolkit compliant; patient data has been pseudonymised for analysis and linkage using industry standard cryptographic hashing techniques; all pseudonymised datasets transmitted for linkage onto OpenSAFELY are encrypted; access to the platform is via a virtual private network (VPN) connection, restricted to a small group of researchers; the researchers hold contracts with NHS England and only access the platform to initiate database queries and statistical models; all database activity is logged; only aggregate statistical outputs leave the platform environment following best practice for anonymisation of results such as statistical disclosure control for low cell counts. The OpenSAFELY research platform adheres to the obligations of the UK General Data Protection Regulation (GDPR) and the Data Protection Act 2018. In March 2020, the Secretary of State for Health and Social Care used powers under the UK Health Service (Control of Patient Information) Regulations 2002 (COPI) to require organisations to process confidential patient information for the purposes of protecting public health, providing healthcare services to the public and monitoring and managing the COVID-19 outbreak and incidents of exposure; this sets aside the requirement for patient consent. Taken together, these provide the legal bases to link patient datasets on the OpenSAFELY platform. GP practices, from which the primary care data are obtained, are required to share relevant health information to support the public health response to the pandemic, and have been informed of the OpenSAFELY analytics platform.

This study was approved by the Health Research Authority (REC reference 20/LO/0651) and by the LSHTM Ethics Board (reference 21863).

This project includes data from the UKRR derived from patient-level information collected by the NHS as part of the care and support of kidney patients. We thank all kidney patients and kidney centres involved. The data are collated, maintained, and quality assured by the UKRR, which is part of the UK Kidney Association. Access to the data was facilitated by the UKRR’s Data Release Group. UKRR data are used within OpenSAFELY to address a limited number of critical audit and service delivery questions related to the impact of COVID-19 on patients with kidney disease.

SRR: Ethical Statement

Formal ethical approval was waived according to Public Health Scotland Information Governance as analysis of routinely collected data. As the analysis used routinely collected and anonymized data, individual patient consent was not sought. Access and use of the data for the purpose of this work were approved following a Public Health Scotland Information Governance review of linking internal datasets. Only the Public Health Scotland analyst had access to the linked patient data, which could only be accessed via an NHS secure network.

## Data access and verification

Access to the underlying identifiable and potentially re-identifiable pseudonymised electronic health record data is tightly governed by various legislative and regulatory frameworks, and restricted by best practice. The data in OpenSAFELY is drawn from General Practice data across England where TPP is the Data Processor. TPP developers (CB, JC, JP, FH, and SH) initiate an automated process to create pseudonymised records in the core OpenSAFELY database, which are copies of key structured data tables in the identifiable records. These are linked onto key external data resources that have also been pseudonymised via SHA-512 one-way hashing of NHS numbers using a shared salt. Bennett Institute for Applied Data Science developers and PIs (BG, CEM, SB, AJW, KW, WJH, HJC, DE, PI, SD, GH, BBC, RMS, ID, TW, TO, SM, CLS, LB, KB, EJW and CTR) holding contracts with NHS England have access to the OpenSAFELY pseudonymised data tables as needed to develop the OpenSAFELY tools. These tools in turn enable researchers with OpenSAFELY Data Access Agreements to write and execute code for data management and data analysis without direct access to the underlying raw pseudonymised patient data, and to review the outputs of this code. All code for the full data management pipeline—from raw data to completed results for this analysis—and for the OpenSAFELY platform as a whole is available for review at github.com/OpenSAFELY.

## Guarantor

DN/LAT are guarantors.

